# Proton pump Inhibitor effect on esophageal protein signature of eosinophilic esophagitis, prediction and evaluation of treatment response

**DOI:** 10.1101/2023.11.21.23298292

**Authors:** Francisca Molina-Jiménez, Lola Ugalde-Triviño, Laura Arias-González, Carlos Relaño-Rupérez, Sergio Casabona, José Andrés Moreno-Monteagudo, María Teresa Pérez-Fernández, Verónica Martín-Domínguez, Jennifer Fernández-Pacheco, Emilio José Laserna-Mendieta, Patricia Muñoz-Hernández, Jorge García-Martínez, Javier Muñoz, Alfredo J Lucendo, Cecilio Santander, Pedro Majano

## Abstract

**Background:** Recently, we have identified a dysregulated protein signature in the esophageal epithelium of eosinophilic esophagitis (EoE) patients; however, the effect of proton pump inhibitor (PPI) treatment on this signature is unknown. Herein, we used a proteomic approach to investigate: (1) whether PPI treatment alters the esophageal epithelium protein profile observed in EoE patients and (2) whether the protein signature at baseline predicts PPI response.

**Methods:** We evaluated the protein signature of esophageal biopsies using a cohort of adult EoE (n=25) patients and healthy controls (C) (n=10). In EoE patients, esophageal biopsies were taken before (Pre) and after (Post) an 8-week PPI treatment, determining the histologic response. Eosinophil count PostPPI was used to classify the patients: ≥15 eosinophils/hpf as non-responders (NR) and <15 eosinophils/hpf as responders (R). Protein signature was determined and differentially accumulated proteins (DAP) were characterized to identify altered biological processes and signaling pathways.

**Results:** High dimensional analysis of DAP between groups revealed common signatures between three groups of patients with inflammation (R-PrePPI, NR-PrePPI and NR-PostPPI) and without inflammation (C and R- PostPPI). PPI therapy almost reversed the EoE specific esophageal protein signature, which is enriched in pathways associated with inflammation and epithelial barrier function, in R-PostPPI. Furthermore, we identified a set of candidate proteins to differentiate R-PrePPI and NR-PrePPI EoE patients before treatment.

**Conclusion:** These findings provide evidence that PPI therapy reverses the alterations in the protein profile associated with EoE. Interestingly, our results also suggest that PPI response could be predicted at baseline in EoE.

## INTRODUCTION

Eosinophilic esophagitis (EoE) is a chronic, immune-mediated inflammatory disease that is characterized by esophageal dysfunction and infiltration of the esophagus by eosinophils ^1–3^. EoE diagnosis is assessed by an endoscopy with esophageal biopsies. Signs and symptoms of chronic or recurrent esophageal dysfunction (e.g., dysphagia, food impaction, heartburn, reflux, chest pain), and histologically, eosinophil infiltration in the esophageal mucosa of over 15 cells per high-power field (hpf) are used for diagnosis^4,5^. Epithelial injury is commonly observed in EoE biopsies, together with hyperplasia of the basal layer, tissue regeneration, and fibrosis in the lamina propria^6^. The natural course of EoE is chronic and apparently progressive, with continuous inflammation that may progress to tissue remodeling thus leading to rigidity and esophageal narrowing in the long term^2,3^.

EoE management includes proton pump inhibitor (PPI), topical steroids, and diet exclusion therapy as first-line therapies^4,5^. Patients with fibrotic strictures or narrow-caliber esophagus should be assessed with endoscopic dilation^7^. Furthermore, high patient relapse rates, and the associated need for long-term therapies have promoted the search of novel EoE treatments including biological therapies targeting type 2 immune responses^8^. PPIs, omeprazole and derivatives, are the most commonly prescribed first-line therapy for EoE because of their efficacy, safety profile, easy administration, and low cost^9^.Their efficacy in EoE has been reported by multiple studies to be in the range of 33% to 50% depending on the criteria used to define histologic remission^10^. In early times, a consensus recommendation for EoE diagnosis postulated trying PPI to exclude gastroesophageal reflux disease (GERD), since this entity can elicit an acid-induced esophageal eosinophilia^11,12^. However, currently, EoE and PPI-responsive esophageal eosinophilia (PPI-REE) are considered as variations of a single disease and a PPI response is no longer considered as a diagnostic criterion in EoE^13^. This consensus has been consolidated since clinical characteristics, endoscopic and histologic findings, pH monitoring, and tissue/genetic markers have failed to distinguish EoE from PPI-REE^9^. In particular, a gene-based EoE diagnostic panel (EDP) has demonstrated that untreated PPI-responders share a largely similar molecular transcriptome with non-responders^14^.

Several studies suggest that the underlying mechanism for the efficacy of PPIs in EoE depends on their anti-inflammatory effect and even on their acid suppression capacities^9^. These dual effects of PPI could contribute to resolution of esophageal eosinophilia in both GERD and EoE. Despite the need for better classification tools to assess PPI response in EoE, there are few data identifying molecular or clinical predictors of PPI response^9^. Shoda et al. established a molecular endotype-based classification in EoE with clinical potential and therapeutic significance, including PPI treatment^15^. Genetic determinants that influence response to PPI such as pharmacogenetic variants in STAT6 and CYP2C19, have been described^16^.

Herein, we hypothesized that the differences observed in protein accumulation between esophageal biopsies from controls and EoE patients^17^ might be altered by PPI-treatment. Furthermore, protein signature analysis could distinguish between responders and non-responders before PPI treatment, helping in the management of EoE patients.

## MATERIALS AND METHODS

### Subjects

Adult EoE patients were prospectively recruited at two Spanish hospitals, Hospital Universitario de La Princesa (Madrid), and Hospital General de Tomelloso (Ciudad Real) between February 2018 and November 2020^17^. EoE was diagnosed according to evidence-based guidelines^4,5^. For EoE diagnosis purposes, three esophageal biopsies were obtained at the distal and proximal esophagus^4,5^. Esophageal eosinophilia was defined as an eosinophil count of ≥15 cells per hpf (corresponding to an area of 0.24 mm2) in one or more biopsy specimens. EoE patients underwent an 8-week course of PPI therapy (20-40 mg of PPI available agents twice daily) and after that, esophageal biopsies were taken. PPI response was defined as positive when the post-treatment esophageal eosinophilia was resolved (R-PostPPi, N=14) (<15 eosinophils/hpf). After histologic evaluation, all NR-PostPPi patients (N=11) exhibited ≥15 eosinophils/hpf at post-treatment. Controls were subjects who underwent upper endoscopy for assessment of dyspepsia or suspected gastroduodenal ulcer. All selected control subjects exhibited a normal endoscopic appearance of the esophagus and they did not meet clinical or histological criteria for EoE after endoscopy and biopsy. Clinical data including demographics, symptoms, atopic background and endoscopic findings were recorded from all EoE patients and control subjects.

This study (PI17/0008) was approved by the Research Ethics Committee of Instituto de Investigación Sanitaria Hospital Universitario de La Princesa (registry number 3107, 8 June 2017). All patients and controls signed an informed consent form before sampling.

### Esophageal biopsies processing, mass spectrometry, RNAseq and bioinformatics and statistical analysis

A detailed description of the methods used has been included in the supplementary Material and Methods section. Information of the antibodies employed is summarized in the **supplementary file 1-Table1.**

## RESULTS

### Characteristics of EoE and control patients

Clinical and demographic characteristics of study participants are summarized in Table 1. Of the 25 EoE patients, 14 (56%) were classified as PPI-responders (R) and 11 (44%) as PPI-non-responders (NR) **(Table 1)**. Compared with controls (n=10) and responders, non-responder patients were older (32 and 35.92 vs 46.63 years respectively), although significant differences were only found between control and non-responders. Non-responders were more frequently male compared to control and responders (100% vs 60% and 78%), but also significant differences were only found between non-responders and the control group. Of note, there were no differences in demographic variables between EoE groups. There were no significant differences regarding symptoms, endoscopic (EREFS score^18^) and histological (EoE-HSS^19^) findings, or PPI treatment options at baseline. The PPI therapy lowered eosinophil count to <15 cells/hpf in all PPI responder cases, reducing the mean peak eosinophil count significantly from 53.64 to 2.28 (p<0.001) **(Table 1).**

**Table 1.** Clinical characteristics of EoE patients before PPI treatment (responders and non-responders) and healthy controls (C). Fisher’s test and Student’s t-test were used to analyze significant demographics and clinical differences between groups, p-value is indicated.

|  | Control<br>(n=10) | Responders<br>(n=14) | Non-responders<br>(n=11) | (C vs R) | *p-value*<br>(C vs NR) | (R vs NR) |
| --- | --- | --- | --- | --- | --- | --- |
| Sex (male) (n,%) | 6 (60%) | 11 (78%) | 11 (100%) | 0.392 | **0.035** | 0.23 |
| Age (mean years ± s.d.) | 32 ±11.6 | 35.92 ± 11.75 | 46.63 ± 14.05 | 0.427 | **0.017** | 0.454 |
| *Symptoms (n,%)* |  |  |  |  |  |  |
| Dysphagia | 0 | 13 (93%) | 11 (100%) | **0.002** | **<0.001** | 1 |
| Food impaction | 0 | 11 (78%) | 7 (63%) | **0.001** | **0.004** | 0.389 |
| Heartburn | 0 | 4 (28%) | 4 (36%) | 0.114 | 0.090 | 1 |
| Abdominal pain | 0 | 0 (0%) | 2 (18%) | 1 | 0.476 | 0.183 |
| *Any atopic disease (n,%)* |  |  |  |  |  |  |
| Asthma | 0 | 7 (50%) | 1 (0.9%) | **0.018** | 1 | **0.042** |
| Allergic rhinitis/ sinusitis | 1 (10%) | 11 (78%) | 10 (90%) | **0.004** | **<0.001** | 0.604 |
| Food allergy | 2 (20%) | 6 (43%) | 1 (0.9%) | 0.387 | 0.586 | 0.09 |
| *Endoscopic findings (n,%)* |  |  |  |  |  |  |
| EREFS (score) (mean ± s.d.) | 0 | 3.5 ± 1.9 | 4.3 ± 1.6 | **<0.001** | **<0.001** | 0.672 |
| Maximum eosinophil count<br>(mean ± s.d.) | 0 | 53.6 ± 31.6 | 61.6 ± 21.4 | **<0.001** | **<0.001** | 0.46 |
| Eosinophil count post PPI<br>(mean ± s.d.) | 0 | 2.28 ± 3.49 | 85.54 ± 66.79 | **0.022** | **0.003** | **0.002** |
| *Histological findings* |  |  |  |  |  |  |
| EoEHSS GRADE score (0-1)<br>(mean ± s.d.) | 0 | 0.48 ± 0.18 | 0.49 ± 0.21 | **<0.001** | **0.001** | 0.866 |
| EoEHSS STAGE score (0-1)<br>(mean ± s.d.) | 0 | 0.45 ± 0.17 | 0.42 ± 0.21 | **<0.001** | **<0.001** | 0.751 |

### Differential proteomic profile in esophageal biopsies of responder and non-responder EoE patients compared to controls

We have recently determined a specific proteomic profile of esophageal biopsies associated to EoE^17^. To address whether PPI-treatment promotes changes in this EoE-associated protein signature, we evaluated the global proteomic profile in esophageal biopsies from both R and NR before and after PPI intake. As illustrated by a principal component analysis (PCA) of the global proteomic profile **(Figure 1A)** clear separation between control and EoE patients prior PPI (including R-PrePPI and NR-PrePPI) treatment was observed across the first principal component, which accounted for 11.98% of total sample variance **(Figure 1A, upper lane).** When the expression profiles were plotted according to the first principal component density, R-PrePPI and NR-PrePPI groups showed comparable distribution patterns **(Figure 1A, lower lane),** thus suggesting that a common EoE protein signature pattern existed, independently of PPI response capacity.

**Figure 1.**
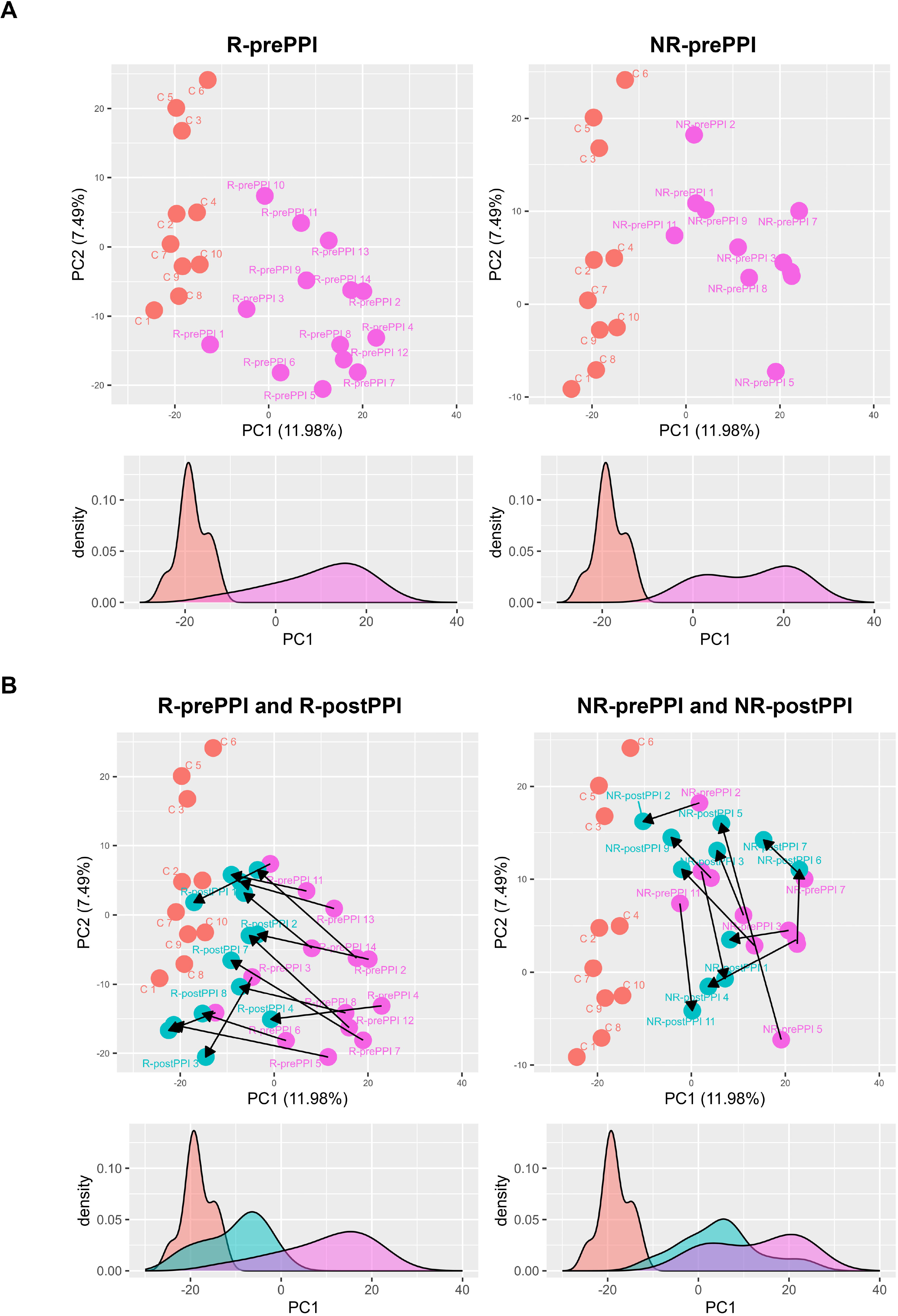
Differential protein expression in esophageal biopsies from EoE patients. A) PCA analysis of whole proteomic data. PC1 and PC2 are represented and the percentage of explained variance is indicated on each axis. The samples are colored by group: A; control (red), R-PrePPI (pink, left), NR-PrePPI (pink, right). B; control (red), R-PrePPI (pink, left), R-PostPPI (green, left), NR-PrePPI (pink, right), NR-PostPPI (green, right). Additionally, PC1 density was plotted (A and B, bottom panels). In paired samples, arrows indicate movement of EoE patients after PPI treatment.

To address whether PPI treatment altered the EoE global protein signature we compared the responders and non-responder cohorts before and after PPI treatment (R-PrePPI vs R-PostPPI and NR-PrePPI vs NR-PostPPI) **(Figure 1B)**. Graphical representation of PCA Principal Components 1 and 2 suggests that PPI therapy is effective at reducing, even eliminating in some patients, differences between R-PostPPI and C associated proteomic profiles, with a clear movement to the left side of the plot. Of note, a modest effect with similar characteristics was also observed in NR-PostPPI patients suggesting that EoE proteomic profile was partially abrogated in non-responder patients, despite their histologic eosinophil-related response was null or incomplete **(Figure 1B).** These results were clearly illustrated when the expression profiles were plotted according to the first component density **(Figure 1B, lower lane)**. Overall, data confirmed that global protein signature from esophageal biopsies could be informative to molecularly define PPI response in EoE patients.

### Effect of PPI treatment on the protein signature of EoE

It has been extensively reported that after PPI therapy, histologic eosinophilia remission is associated with a specific mRNA-signature^14^, but it is unknown whether EoE-associated protein signature recover control values after PPI therapy. To address this, we evaluated DAP in esophageal biopsies from EoE patients before and after PPI treatment. DAP in esophageal biopsies were identified after comparison between control individuals and all groups of EoE patients **(Figure 2A and Supplementary File 1. Tables 2-5)**. Differential analysis (adjusted p-value≤0.05 and fold change>1.5) showed the following results: R-PrePPI vs C (312 DAP, 148 upregulated, 164 downregulated); R-PostPPI vs C (44 DAP, 17 upregulated, 27 downregulated); NR-PrePPI vs C (279 DAP, 138 upregulated, 141 downregulated); NR-PostPPI vs C (144 DAP, 64 upregulated, 80 downregulated proteins).

**Figure 2.**
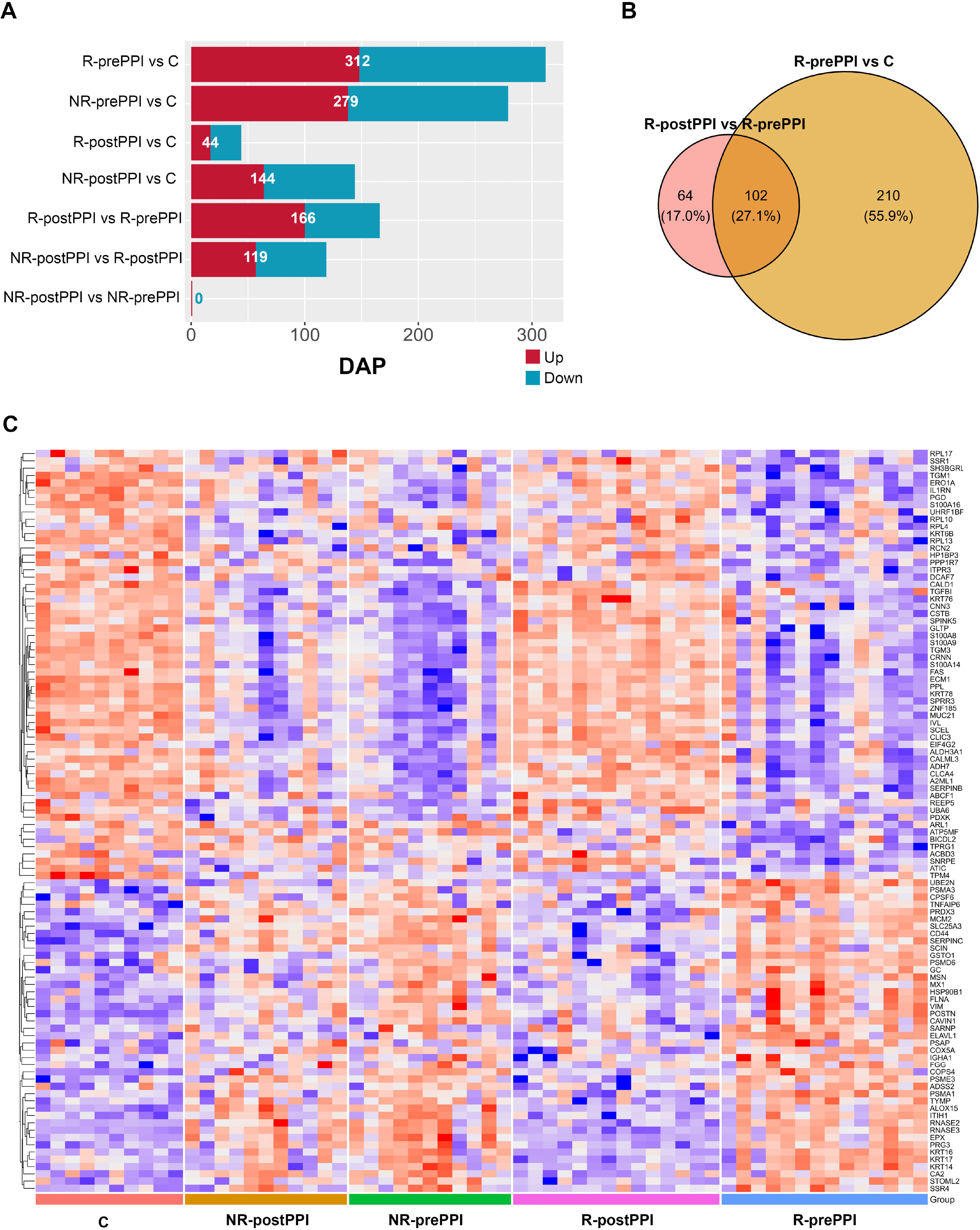
Differential protein expression profiles of study cohorts. A. DAP proteins (adjusted p-value≤ 0.05 and a 1.5-fold change) in esophageal biopsies obtained by comparisons of control and EoE cohorts (C n=10; R-PrePPI n=14; NR-PrePPI n=11; R-PostPPI n=14; NR-PostPPI n=11). Red indicates upregulated DAP, and blue, downregulated DAP relative to the second group in the comparison. Number of DAP is indicated. B. Venn diagrams showing the overlap between dysregulated proteins in both C vs R-PrePPI, and R-PrePPI vs R-PostPPI C. Heatmap of the 102 proteins selected in panel B in the 5 cohorts. z-score indicating protein levels is presented. Red indicates higher expression (upregulation), and blue represents lower expression (downregulation).

Next, we compared responder patients before and after treatment R-PostPPI vs R-PrePPI (166 DAP, 100 upregulated and 66 downregulated) and non-responders NR-PostPPI vs NR-PrePPI (none DAP). Finally, NR-PostPPI were compared to R-Post-PPI (119 DAP, 57 upregulated and 62 downregulated). A complete information regarding DAP obtained in all comparisons is included in **Supplementary File 1-Tables 6-8**.

To identify a protein signature associated with PPI response, common DAP in C vs R-PrePPI and R-PrePPI vs R-PostPPI comparisons were analyzed **(Figure 2B and Supplementary File 1-Table 2 and 6).** Results showed 102 DAP shared by both comparisons **(Supplementary file 1-Table 9)**. Next, we evaluated the expression profile of these 102 DAP in all five groups **(Figure 2C).** As illustrated by the supervised clustered heatmap, there was a remarkable conservation of the esophageal epithelium proteomic profile between C and R-PostPPI groups **(Figure 2C).** In contrast, this pattern was different in the pretreatment groups (R-PrePPI, NR-PrePPI) and post-treatment NR-PostPPI group. R-PostPPI samples showed similar levels to control samples probably associated with the lower inflammatory state of these patients. In contrast, groups with esophageal eosinophilia (R-PrePPI, NR-PrePPI and NR-PostPPI) showed more similar patterns between them but different to control and R-PrePPI patients. This protein panel associated to PPI-response included eosinophil related proteins not detected at mRNA level (ribonuclease A family member 2, RNAse2; ribonuclease A family member 3, RNAse3; eosinophil peroxidase, EPX), and 32 esophagus enriched genes ^20^ **(Supplementary File 1-Table 9)** strongly suggesting that altered pathways in EoE^17^ related to inflammation and epithelial differentiation are recovered in R-PostPPI patients.

### Immunofluorescence-based assessment of protein expression confirms proteomic data

Next, we validated the expression of several DAP that are representative of the inflammatory and epithelial alterations observed between control and the different EoE cohorts by immunofluorescence in esophageal biopsies. We selected two proteins that were significantly upregulated in EoE patients with inflammation (R-PrePPI, NR-PrePPI and NR-PostPPI) comparing them to those without inflammation (C, R-PostPPI) (CD44; Arachidonate 15-Lipoxygenase, ALOX15). In addition, we analized two proteins that were significantly downregulated (cornulin, CRNN; mucin21, MUC21) in inflamed samples. Immunofluorescence analysis showed similar results to proteomic data highlighting that an effective PPI treatment reverse the EoE-associated protein signature **(Figure 3).**

**Figure 3.**
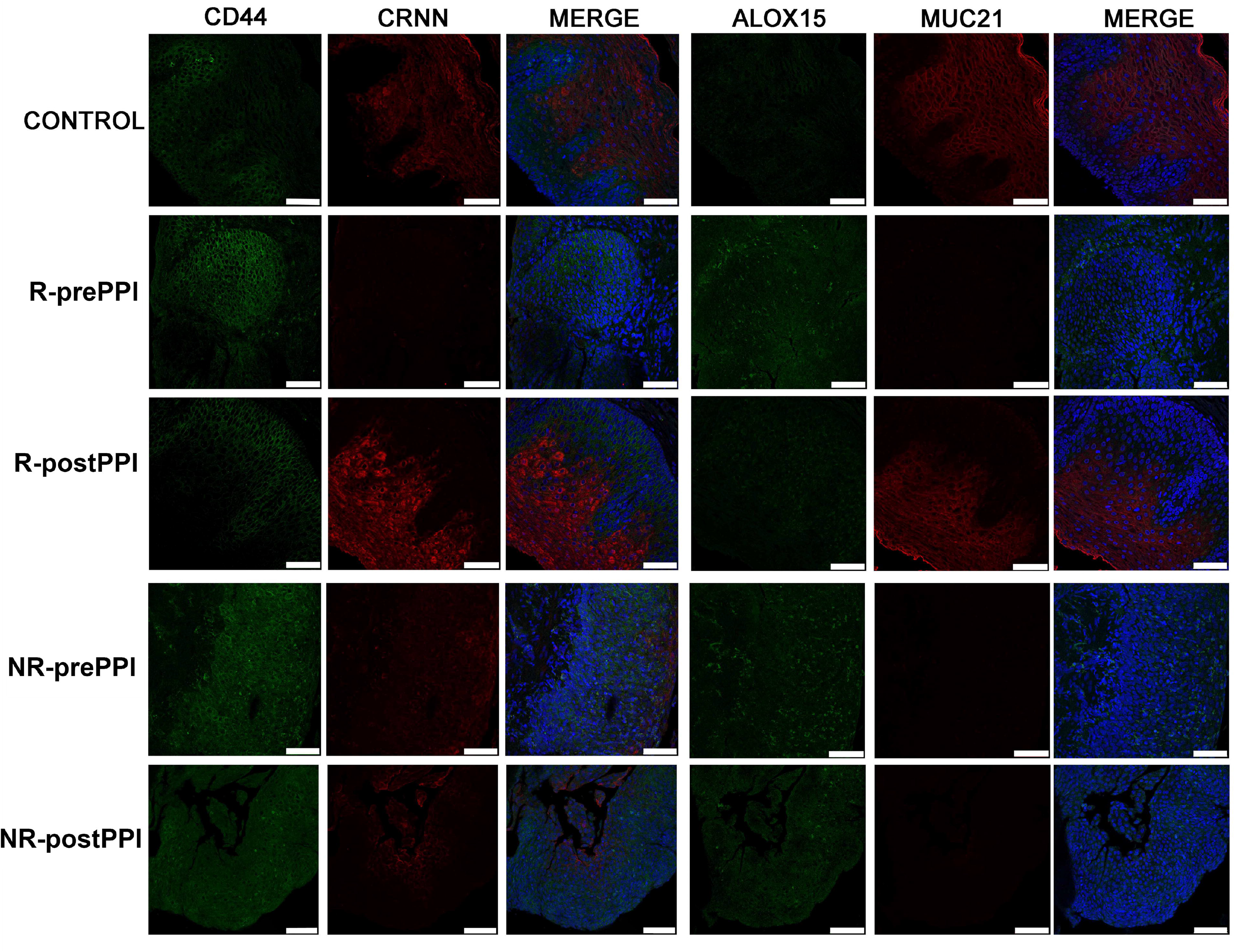
Expression of selected DAP in esophageal biopsies from PPI responder EoE patients. Immunofluorescence analysis of CD44, CRNN, ALOX15 and MUC21 in esophageal biopsy sections from a presentative control subject and representative R-PrePPI, R-PostPPI, NR-PrePPI and NR-PostPPI EoE patients. Serial tissue sections were stained with pairs of antibodies to simultaneously detect: CD44 (green) and CRNN (red); ALOX15 (green) and MUC21 (red). DAPI (blue) is shown in merge images. Scale bar 75_Jμm. Images are representative of each group.

### In depth protein signature analysis after PPI treatment

To thoroughly characterize the effect of PPI on esophageal biopsies, we evaluated the protein profile in paired samples from responder patients before and after treatment with PPI. In this cohort, eight weeks of PPI therapy reduced the mean peak eosinophil count significantly from 53.64 to 2.28 (p<0.001), as well as the EREFS score, from 3.5 to 1.41 (p<0.001) and the EoE HSS score from 0.48 to 0.04 (p<0.001) **(Table 2)**. On the other hand, non-responding patients did not present any significant changes in their eosinophil peak count and histological and endoscopic scores when compared to baseline **(Table 2).**

**Table 2.** Clinical characteristics of EoE responder and non-responder patients before and after PPI treatment. Fisher’s test and Student’s t-test were used to analyze significant clinical differences between groups, p-value is indicated.

|  | **R-prePPI**<br>**(n=14)** | **R-postPPI**<br>**(n=14)** | **p-value** |
| --- | --- | --- | --- |
| *Endoscopic findings (n,%)* |  |  |  |
| EREFS (score) (mean ± s.d.) | 3.5 ± 1.91 | 1.41 ± 1.29 | <0.001 |
| Maximum eosinophil count<br>(mean ± s.d.) | 53.64 ± 31.57 | 2.28 ± 3.49 | <0.001 |
| *Histological findings* |  |  |  |
| EoEHSS GRADE score (0-1)<br>(mean ± s.d.) | 0.48 ± 0.18 | 0.04 ± 0.04 | <0.001 |
| EoEHSS STAGE score (0-1)<br>(mean ± s.d.) | 0.45 ± 0.17 | 0.01 ± 0.03 | <0.001 |
|  | **NR-prePPI**<br>**(n=11)** | **NR-postPPI**<br>**(n=11)** | **p-value** |
| *Endoscopic findings (n,%)* |  |  |  |
| EREFS (score)(0-9) (mean ± s.d.) | 4.27 ± 1.61 | 4 ± 2.28 | 0.75 |
| Maximum eosinophil count<br>(mean ± s.d.) | 61.63 ± 21.4 | 85.54 ± 66.8 | 0.284 |
| *Histological findings* |  |  |  |
| EoEHSS GRADE score (0-1)<br>(mean ± s.d.) | 0.49 ± 0.21 | 0.37 ± 0.21 | 0.184 |
| EoEHSS STAGE score (0-1)<br>(mean ± s.d.) | 0.42 ± 0.21 | 0.31 ± 0.21 | 0.227 |

A total of 166 proteins were identified as DAP when comparing responder patients before and after treatment **(Figure 4A, left).** Specifically, 100 proteins were upregulated, and 66 were downregulated **(Supplementary File 1 Table 6).** In addition, a heatmap was generated using the DAP from the previous analysis **(Figure 4B)** clearly showing a different protein accumulation profile. On the contrary, non-responding patients when compared before and after treatment did not present a substantial variation in their proteomic profile and no DAP were found **(Figure 2A).**

**Figure 4.**
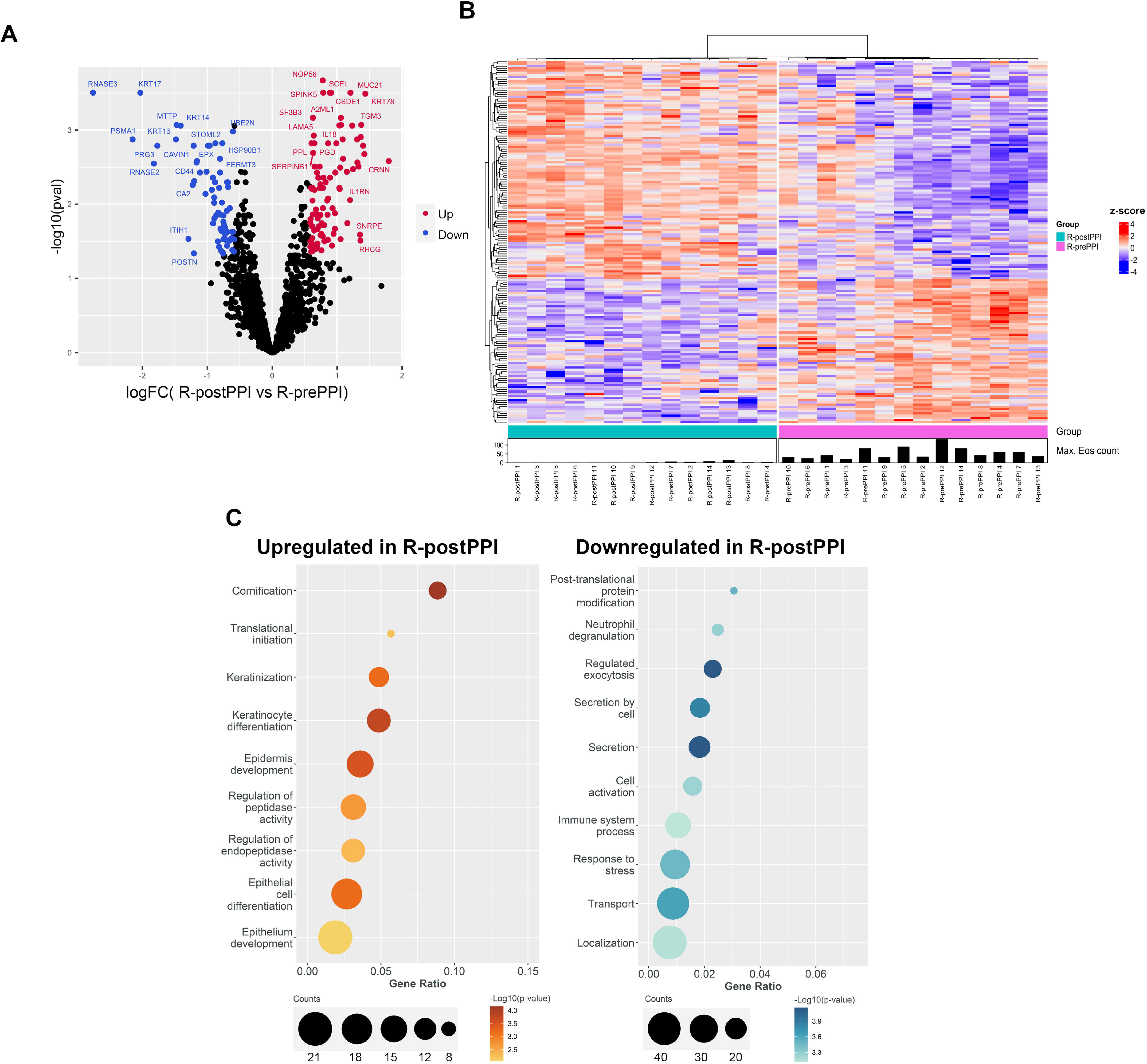
Differential protein expression in PPI response. A) Volcano plot representation of the differential expression analysis. Log2 fold change is represented on the X axis, and -log10 of adjusted p-values on the Y axis. The proteins are colored by relative expression values: upregulated or downregulated (red and blue, respectively) in R-PostPPI.. Differential expression criteria are an adjusted p-value ≤ 0.05 and a 1.5-fold change. Gene names are shown for the most extreme values. (B) Heatmap showing Z-score scaled protein expression of 166 differentially expressed proteins between R-PostPPI and R-PrePPI patients. Sample group is indicated, with the maximum eosinophil count from each subject. The proteins are hierarchically clustered using Euclidean distance, as shown in the top dendrogram (C) Gene enrichment analysis of DAP in PPI responder patients. Representation of the most relevant Gene Ontology (GO) terms related to biological processes. The size of the dot represents the number of proteins from our data set related to each process. Dots are colored according to their significance, which is set by a color scale referring to -log10 (adjusted p-value). Left (upregulated in R-PostPPI), right (downregulated in R-PostPPI).

To identify biological processes and pathways in which the DAP (with p-value≤0.05 logFC>1.5) in PPI-responder patients are involved, we performed a gene enrichment analysis using String software tool (https://string-db.org/)^21^,using adjusted p-value ≤ 0.05. Immune activation related pathways, including neutrophil degranulation, were significantly enriched in upregulated proteins in responding PrePPI patients (adjusted p-value<10^-11^) **(Figure 4C)**. Proteins associated with vesicle mediated transport were also upregulated, suggesting an active vesicle trafficking in the damaged areas. In the downregulated protein subset, we identified cornification and keratinization as the most enriched biological processes (adjusted p-value<10^-9^ and <10^-14^, respectively) **(Figure 4C).**

Remarkably, we found that several DAP between R-PostPPI and R-PrePPI were undetectable at mRNA level in our previous analysis **(Supplementary File 1-Table 10).** Applying a selective criterion (fold change>1.5 adjusted p-value≤0.05) and focusing on the most relevant proteins in EoE three main groups of DAP could be observed **(Supplementary File1-Table 10)**. The first includes four eosinophil granule-derived proteins, proteoglycan 3 (PRG3), ribonuclease A family member 3 (RNASE3), eosinophil peroxidase (EPX), and RNASE2. A second group including Inter-alpha-trypsin inhibitor heavy chain (ITIH1), serpin family C member 1(SERPINC1), and microsomal triglyceride transfer protein (MTTP) are typically synthesized in the liver with a constitutive secretion into blood^20^. The third, a miscellaneous group, includes a keratin (KRTN-76) and myosin heavy chain 7B (MYH7), which are also expressed by basophils, according to the Human Protein Atlas database^20^ and were previously detected in the esophageal mucosa by a global human tissue proteomic iniciative^22^.

It has been described that 39% of the esophagus-specific transcripts (117) are altered in esophageal biopsies from EoE patients (Eso-EoE panel) being around 90% of them downregulated^23^. Using the current data available at the Human Protein Atlas site^20^, we found that 40 DAP from the PPI-responder comparison were esophagus-enriched genes, of which 38 were upregulated and only 2 downregulated in R-PostPPI vs R-PrePPI **(Supplementary** Figure 1 **and Supplementary File 1-Table 10)**. Overall, these data suggested that our proteomic analysis agreed with Eso-EoE panel^23^ confirming a downregulation of esophagus enriched genes and a profound loss of proteins related with esophageal epithelial differentiation in EoE.

### Prediction of PPI response in EoE: Protein signature in PPI responders vs non-responders at baseline

The identification of molecular factors that might predict response to PPI in patients with EoE has been largely elusive^9^. In our cohort, there were no significant differences between responders and non-responder patients at baseline in demographics, symptoms, and endoscopic and histologic findings **(Table 1).**

To further assess whether protein signatures at baseline differed, protein profiles from esophageal biopsies were compared between R-PrePPI and NR-PrePPI EoE. First, the global transcriptomic profile was analyzed by PCA showing no differences in the transcriptomic signature between responder and non-responder patients across both the first and second principal component, which accounted for 31.95% and 10.06% of total sample variance, respectively (**Figure 5A**). Our transcriptomic analysis revealed that no DEG between groups were detected at baseline (adjusted p-value≤0.05 and fold change>2) **(supplementary File1-Table 12).** These results agreed with previously reported studies that failed to find any molecular difference between these two groups^9^. Interestingly, when this analysis was performed using proteomic data, PCA results showed a clear separation between R-PrePPI and NR-PrePPI patients across the second principal component, which accounted for 7.4% of total sample variance **(Figure 5B)**. Further differential analysis of protein accumulation between these two cohorts identified 28 DAP (2% of total detected proteins) (adjusted p-value≤0.05 and fold change>1.5) **(Figure 5C)**. Specifically, 12 proteins were upregulated and 16 were downregulated in NR-PrePPI when compared to R-PrePPI **(Figure 5 and Supplementary File1-Table 11)**. In addition, a supervised heatmap was generated using the DAP from the previous analysis showing a clear difference in the pattern of protein accumulation **(Figure 5D).** To identify the over-represented GO terms (biological processes) associated with the DAP (with p-value≤0.05 logFC > 1.5) between responders and non-responders PrePPI patients, we performed gene enrichment analysis. Interestingly, NR-PPI patients presented increased levels of proteins related with regulated exocytosis and antigen presentation processes **(Figure 5E)**. Finally, to further confirm these observations we validated the expression of two selected DAP by immunofluorescence analyses in representative tissue samples (R-PrePPI and NR-PrePPI EoE patients): Histocompatibility Minor 13, HM13 and Protein Phosphatase Methylesterase 1, PPME1 **(supplementary Figure 2).**

**Figure 5.**
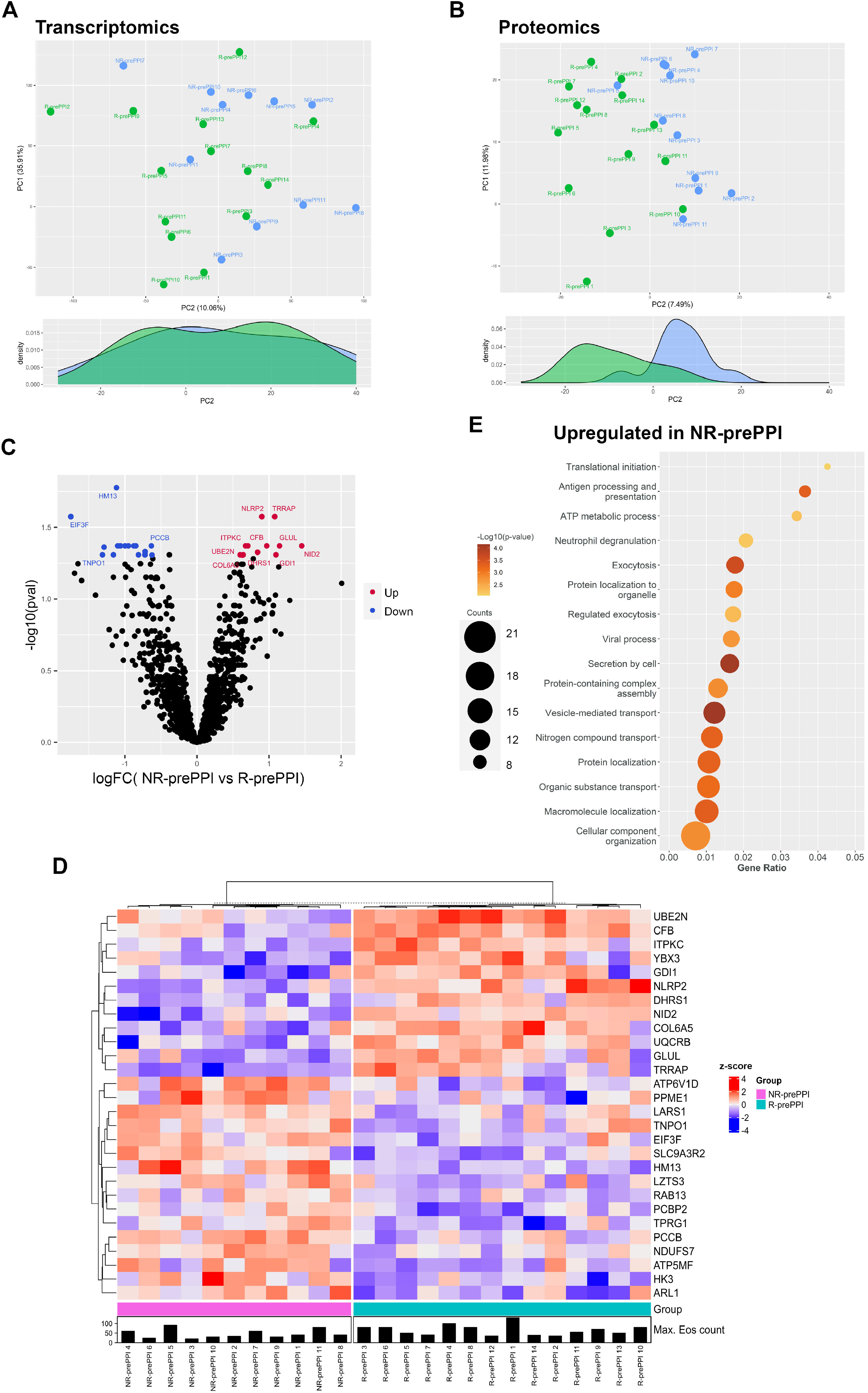
Differential protein expression in responder and non-responder patients at baseline. A) Representation of PCA of whole transcriptome data, PC1 on y axis and PC2 on x axis. Percentage of explained variance is indicated on each axis. The samples are colored by group; R-Pre-PPI (green), NR-PrePPI (blue). B) PCA representation of proteomic data, PC1 on y axis and PC2 on x axis. Percentage of explained variance is indicated on each axis. The samples are colored following the same key as in A. C) Volcano plot representation of the differential expression analysis in the two Pre-PPI EoE cohorts. Log2 fold change is represented on the x axis, and -log10 of adjusted p-values on the y axis. The proteins are colored by relative expression values: upregulated or downregulated in NR-PrePPI (red and blue, respectively). Differential expression criteria are adjusted p-value ≤ 0.05 and a 1.5-fold change. Gene names are shown for the most extreme values. (D) Heatmap showing z-score scaled protein expression from 28 differentially expressed proteins between R-PrePPI and NR-PrePPI EoE patients. Sample group is indicated on the bottom bar, with the maximum eosinophil count from each subject. The proteins are hierarchically clustered using Euclidean distance, as shown in the left dendrogram. (E) Gene enrichment analysis of differentially expressed proteins NR-PrePPI vs R-PrePPI. Representation of the most relevant Gene Ontology (GO) terms related to biological processes. The size of the dot represents the number of genes from our data set related to each process. Dots are colored according to their significance, which is set by a color scale referring to -log10 (adjusted p-value).

In summary, our analyses showed no substantial differences in baseline mRNA levels between EoE patients who will respond to PPI therapy and those who will not. However, we found a specific protein signature able to explain a percentage of the variability between responding and non-responding patients before PPI treatment.

## DISCUSSION

Herein, we used an unbiased proteomic approach to provide, for the first time, an expanded view of the molecular changes occurring within the inflamed esophageal mucosa of EoE patients treated with PPI. We found that untreated EoE patients at baseline, including PPI responders and non-responders, have a similar altered protein signature in the esophageal tissue when compared to control individuals **(Figure 1)**, providing compelling evidence that the two groups are part of the same disease^24^. Nonetheless, the resulting differentially accumulated protein panel **(Figure 2C),** which includes proteins involved in inflammatory process and epithelial differentiation, was almost reverted after PPI treatment in EoE responder patients **(Figures 1-4)**. We confirmed that the EoE esophageal proteome reflects the esophageal tissue alteration, associated with a chronic eosinophilic inflammatory condition, reversed by an effective PPI treatment. Recent studies support differential symptomatic and histopathological features for adult and pediatric EoE patients, but a common pathogenesis and similar response to treatment^25^. Whether this EoE-associated protein signature observed in adults could be also applied to children remains to be determined.

It has been previously reported that some EoE patients have a decline in eosinophil count after PPI treatment without achieving histologic response, whereas in other cases histologic parameters such as lamina propria fibrosis, eosinophil degranulation, and basal cell hyperplasia also decrease^26–28^. Interestingly, although no DAP were found between NR-PrePPi and NR-Post PPI (Figure 2 and supplementary File1-Table 7), a partial recovery of the protein signature was also observed in non-responder patients **(Figure 1).** More in-depth analysis of the non-responder group revealed that 4 patients (36.3%) achieved partial reduction in the eosinophil count after PPI treatment, with 2 patients (18.2%) having ≥50% decrease. In particular, several NR-PostPPI patients showed a clear shift towards the protein profile observed in control or R-PostPPI patients **(Figure1)**. In these patients the histologic index (EoE-HSS) before and after PPI treatment was substantially reduced, regardless of the eosinophil count. These findings raise the question of whether there is a subgroup of EoE patients who may benefit from ongoing PPI treatment, even in the absence of a histologic response^29^.

Around 70-80% of those EoE patients in whom high PPI doses achieve histological and clinical remission maintain long-term remission after a dose reduction^29,30^. Our results demonstrate that short-term PPI treatment restores the accumulation of proteins related to inflammation and epithelial differentiation processes such as cornification **(Figure 4)**, suggesting that it might also prevent long-term complications of EoE such as scarring and narrowing of the esophagus^28^. However, whether PPI maintenance therapy, usually combined with a dose reduction, preserves this non-inflamed protein signature needs to be determined.

In patients with clinical and histologic features of EoE, genotypic and phenotypic features of PPI responders and non-responders are indistinguishable^9^. Unfortunately, PPI have short- and long-term adverse side effects making unnecessary or too long exposure risky for patients^9^. Therefore, clinical, symptomatic and/or molecular approaches to identify PPI response before intervention would be an important advance for patient care. In our work, we identified a specific protein profile **(Figure 5)** capable of predicting PPI response before starting treatment. When we categorized DAP between R and NR PrePPI, they were involved in two main signaling pathways related with regulated exocitosis and antigen presentation **(Figure 5)**. Considering the upregulation of antigen presentation at tissue level **(Figure 5)**, our observations could be indicating a high infiltration of these cells in the esophageal tissue during the sustained inflammation. These patients might present a more altered barrier in the esophagus, increasing the risk of antigen infiltration, thereby favoring EoE worsening, as described before^31^. In the case of topical corticosteroids, inflammatory mediators have been proposed as predictors of treatment response^32^. Our results described a protein signature at baseline able to identify NR-PPI patients that could help clinicians with decisions on best treatment options. Nevertheless, further analysis is needed to validate whether a protein signature could be used as a predictor for best therapeutic option decisions in routine clinical practice, even for other first-line therapies in EoE such as topical corticosteroids or elimination diets.

Current recommendations for monitoring the therapeutic effect after any treatment in EoE involve serial upper gastrointestinal tract endoscopy with biopsies^4,5^. This procedure entails a risk for complications and requires sedation. Identifying non-invasive or minimally invasive biomarkers is not only of high interest for treatment response prediction but also for diagnosis and patient follow-up^33^. Interestingly, since the protein signature is closer to clinical features, our work could lay the groundwork for novel diagnostic biomarkers. Despite our proteomic approach was carried out in esophageal biopsies, according to the Uniprot database, 29 of 102 proteins defined as indicative of patient recovery are classified as secreted proteins (GOTerm GO:0005576, extracellular-region) **(supplementary File1-Table 9),** while 32 are classified as esophagus enriched genes. Furthermore, 4 of 28 proteins defined as indicative of PPI response are classified as secreted proteins, and 3 are classified as esophagus enriched genes **(supplementary File1-Table 11)**. Overall, these proteins could be theoretically measured in blood or luminal esophageal samples obtained by minimally invasive methods (as the string test^34^or cytosponge devices^35^).

In summary, we have described for the first time that the protein signature of the allergic inflammation associated with active EoE was reverted after PPI treatment in responder patients. Furthermore, despite the protein signature is similar and mRNA signatures are indistinguishable between PPI responding and non-responding patients at baseline, differential protein accumulation between them sugggest new potential non-invasive biomarker predictors.

## Finantial support

PM and CS are supported by grants PI17/0008 and ISCIII-Proteored 2019 of Instituto de Salud Carlos III (ISCIII, Spain) and co-funded by Fondo Europeo de Desarrollo Regional (FEDER). CS is also funded by Asociación Española de Gastroenterología (AEG) 2019 grant. JM is supported by grant PID2021-123144OB-I00 funded by MCIN /AEI/10.13039/501100011033 / FEDER, UE. LU-T is recipient of an INVESTIGO contract from Comunidad de Madrid (09-PIN1-00015.6/2022) partly funded by the European Social Fund, NextGenerationEU, and Recovery, Transformation and Resilience Plan. CR-R is recipient of an INVESTIGO contract from Ministry of Labour and Social Economy, the national public employment service (SEPE) (INVESTIGO Exp. 2022- C23.I01.P03. S0020-0000031) partly funded by the European Social Fund, NextGenerationEU, and Recovery, Transformation and Resilience Plan. EJL-M holds a Juan Rodés grant (JR19/00005) from the ISCIII, Spanish Ministry of Health - Social Services and Equality, which is partly funded by the European Social Fund (2014-2020).

## Data availability statement

The data that supports the findings of this study are available in the supplementary material of this article. Additional dataset will be available upon request at time on publication. Please contact the corresponding author for any inquiries.

## Acknowledgments

We express our gratitude to Drs. Félix Elortza and Mikel Azkargorta (Proteomic Unit, CIC-Biogune) and Dr Manuel José Gómez (Bioinformatic Unit, Centro Nacional de Investigaciones Cardiovasculares-CNIC) for technical support and data analysis. We thank Dr. Manuel Gómez for critical review of the manuscript.

## Author disclosure

None to declare.

## Author contributions

Conceived and designed the study: FM-J, AJL, CS, PM.

Participated in the clinical management of patients: SC, JAM-M, MTF-P, VM-D, JF-P, PM-H, AJL, CS.

Performed the experiments: FM-J, LU-T, LA-G, EJL-M, CR-R.

Analyzed and discussed the data: FM-J, LU-T, CR-R, JG-M, JM, AJL, CS, PM. Wrote the paper: FM-J, LU-T, PM.

All the authors read, provided comments, and approved the final version of the manuscript.

## Abbreviations

C: Control
DAP: Differentially accumulated proteins.
DEG: Differentially expressed genes
EDP: EoE diagnostic panel
EoE: Eosinophilic esophagitis
GERD: Gastroesophageal reflux disease
GO: Gene ontology (GO) terms
hpf: High-power field
IPA: Ingenuity Pathway Analysis
NR-PostPPi: Patient with EoE Non-Responder to PPI post-treatment
NR-PrePPi: Patient with EoE Non-Responder to PPI pre-treatment
PCA: Principal component analysis
PPI: Proton pump inhibitors
PPI-REE: Proton pump inhibitor-responsive esophageal eosinophilia
RNAseq: RNA sequencing
R-PostPPi: Patient with EoE Responder to PPI post-treatment
R-PrePPi: Patient with EoE Responder to PPI pre-treatment

## Notes

### Competing Interest Statement

The authors have declared no competing interest.

### Author Declarations

This study was approved by the Research Ethics Committee of Instituto de Investigacion Sanitaria Hospital Universitario de La Princesa (registry number 3107, 8 June 2017). All patients and controls signed an informed consent form before sampling. All necessary patient/participant consent has been obtained. Any patient/participant/sample identifiers cannot be used to identify individuals.

