## Supplementary Materials and methods for "Proton pump Inhibitor effect on esophageal protein signature of eosinophilic esophagitis, prediction and evaluation of treatment response"

**SUPPLEMENTARY MATERIAL AND METHODS.**

**Esophageal biopsies collection and processing.**

Esophageal biopsies for both clinical and research purposes were taken during the same endoscopic procedure in the proximal and distal esophageal sections. Samples were collected from affected areas if applicable. Proximal and distal samples used for molecular studies were mixed and processed together as previously described**^1^**. Briefly, under liquid N2 freezing conditions, biopsies were disrupted using a mortar and pestle, grinding them to a fine powder. The tissue powder was divided in two parts to extract RNA and proteins.

**Endoscopic and histologic scores.**

The EoE endoscopic reference score (EREFS)^2^ rating the severity of esophageal inflammation (edema, furrows, exudate) and fibrostenosis (rings and stricture) was scored in all patients. Furthermore, the validated Eosinophilic Esophagitis Histologic Scoring System (EoEHSS)^3^ was determined, evaluating eight pathologic features for both severity (grade) and extent (stage) of abnormalities. One of the EoEHSS features (thickened connective tissue in the lamina propria) was excluded because a large proportion (60%) of samples lacked lamina propria^3,4^**.** Moreover, an analysis with Fisher’s Exact Test determined that there was no significant association between patient groups and lamina propria presence.

**Protein extraction and digestion.**

Disrupted tissues were incubated in 7M urea, 2M Thiourea, 4% CHAPS and 5mM DTT. Protein was digested following the filter-aided FASP protocol^5^.

**Mass spectrometry & bioinformatic analysis.**

Samples were analyzed in a novel hybrid trapped ion mobility spectrometry – quadrupole time of flight mass spectrometer (timsTOF Pro with PASEF, Bruker Daltonics) coupled online to an Evosep One LC system. Samples (200 ng each) were directly loaded in an Evosep 8 cm analytical column and resolved using a 21 min gradient. Protein identification and quantification was carried out using PEAKS software (Bioinformatics solutions). Searches were analyzed against a human database (Uniprot 2020_04, 20375 entries), with precursor and fragment tolerances of 20 ppm and 0.05 Da. Two missed cleavages were allowed. Met (Ox)-variable and Cys (Carabamidomethyl)-fixed were considered as modifications. A decoy search was conducted in order to estimate the FDR of the searches – only peptides and proteins identified with an FDR<1% were kept. For the quantitative analysis, only proteins identified with at least two peptides at FDR<1% were considered for further analysis~~.~~ Data were loaded onto the Prostar platform ^6^ and further processed (log2 transformation, filtering, imputation). A Limma paired or unpairedwas applied in order to determine the statistical significance of the differences detected^7^. R software 4.1.3 was used for further analysis and graph generation.

**Immunohistofluorescence staining in esophageal biopsies.**

Immunofluorescence staining was performed with biopsies from control subjects and patients with active EoE prior and after PPi treatment, obtained from distal esophagus, previously oriented in a cellulose acetate and included in paraffin-embedded blocks. Slides with 4-μm sections of FFPE esophageal biopsies, underwent deparaffinization, antigen retrieval using sodium citrate buffer (10 mM sodium citrate, 0.05% Tween 20, pH 6.0), blocked with 10% goat serum/phosphate-buffered saline (PBS), and then incubated with primary antibody diluted in 10% goat serum/PBS overnight at 4 °C in a humidified chamber. The next day, slides were washed with PBS, incubated with secondary antibodies and DAPI (0,5 μg/mL) diluted in 10% goat serum/PBS for 1h at room temperature in a humidified chamber, and then washed with PBS. Finally, a cover slip was added with ProLong Gold mounting reagent (Molecular Probes).

Images were obtained using a Leica TCS SP5 confocal microscope. Immunofluorescence experiments were carried our using the antibodies listed in Supplementary File 1-Table 1.

**GO enrichment analysis.**

The “STRING analysis tool v11.5^8^ was used to perform these analyses.

**RNA-Seq library preparation and sequencing.**

The total RNA samples were quality controlled by Qubit® RNA BR Assay kit (Thermo Fisher Scientific) for quantity and RNA 6000 Nano Assay (Agilent) for integrity. The RNA-Seq libraries were prepared from total RNA using the TruSeq Stranded mRNA Library Prep Kit (Illumina) according to the manufacturer’s guide. The libraries were sequenced on HiSeq4000 (Illumina, Inc) in a fraction of a HiSeq 4000 PE Cluster kit sequencing flow cell lane, following the manufacturer’s protocol for dual indexing. Image analysis, base calling and quality scoring of the run were processed using the manufacturer’s software Real Time Analysis (RTA 2.7.7) and followed by generation of FASTQ sequence files. Read quality was determined with FASTQC and adapter sequences were trimmed with cutadapt v1.14^9^. RSEM v1.3.1^10^ was used to map reads against the reference human genome (GRCh38_p9) and to calculate count matrices. Ensembl gene IDs were used to map transcripts against the human genome. Differential expression analysis was performed using the Limma test in R ^(^https://bioconductor.org/packages/release/bioc/html/limma.html). Only those differentially expressed genes (DEG) with an adjusted p-value of 0.05 (Benjamini-Hochberg correction) were evaluated.

**Statistical analysis.**

Features of the study subjects were summarized with descriptive statistics. Baseline data for cases and controls were compared using two-sample t-test for continuous variables and Fisher’s test for categorical variables. Differences between baseline and post-treatment endoscopic and histologic scores for EoE cases were analyzed using paired t-tests. Proteins were normalized using the TMM method. Limma was performed for protein differential analysis between groups, a paired approach was used for responders and non-responders prior and after PPI treatment, or unpaired for the remaining comparisons^7^. edgeR was used for differential analysis of the RNAseq data^11^**.** For each pairwise comparison, upregulated and downregulated proteins were defined as those with at least a 1.5- fold and adjusted p-value≤0.05. For mRNA a 2.0-fold change and adjusted p-value≤0.05 in expression were used as significant threshold.

**Human Protein Atlas database.**

Definitions related to specificity of expression in the esophagus were obtained from the Human Protein Atlas (<http://www.proteinatlas.org/humanproteome/esophagus>) (Accessed march 2023). Post acceptance updates to the Human Protein Atlas may alter the number of tissue-specific (and esophagus-specific) genes from values presented herein.
