## Supplementary Figure1 for "Proton pump Inhibitor effect on esophageal protein signature of eosinophilic esophagitis, prediction and evaluation of treatment response"

**A**

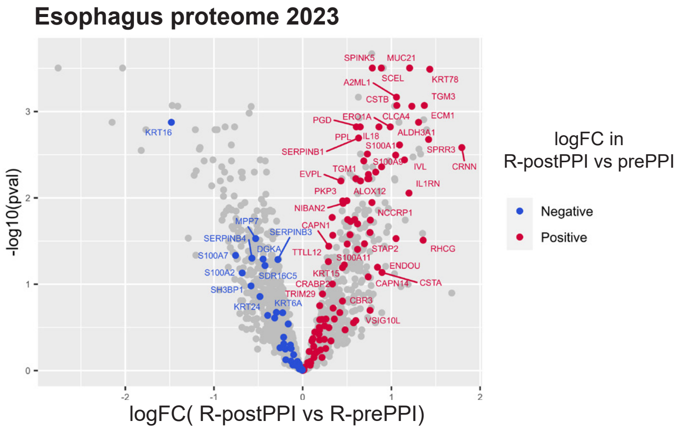


**Supplementary Figure 1:** Comparative study of proteomics analysis in responder patients. Volcano plot of protein differential expression analysis showing the position of esophagus-enriched proteins according to Human Protein Atlas 2023. Dot color indicates the direction of the value of the fold change in the differential analysis of our proteomic data.
