## Supplementary Figure1 for "Proton pump Inhibitor effect on esophageal protein signature of eosinophilic esophagitis, prediction and evaluation of treatment response"

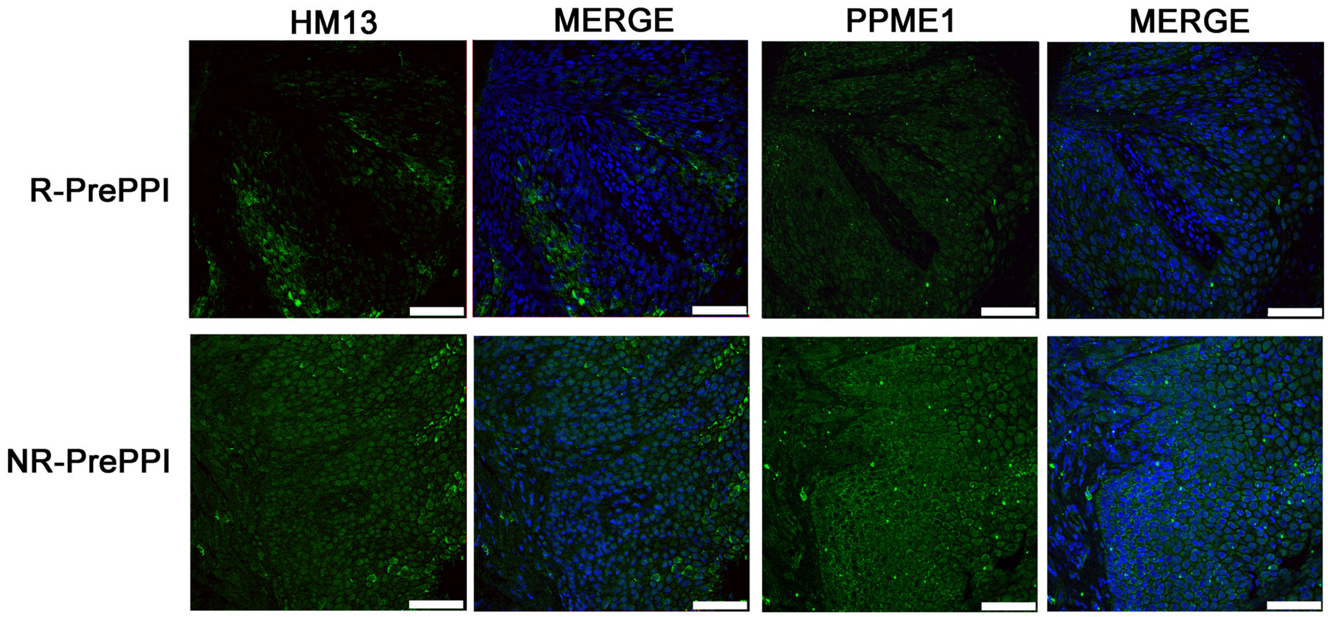


**Supplementary Figure 2.** Expression of selected dysregulated proteins at baseline, R-PrePPI vs NR-PrePPI EoE patients. Immunofluorescence analysis of HM13 and PPME1 (green) in esophageal serial biopsy sections from representative R-PrePPI and NR-PrePPI patients. DAPI (blue) is shown in MERGE images. Scale bar 75 μm.
